# Perils of the nighttime: impact of behavioral timing and preference on mental and physical health in 73,888 community-dwelling adults

**DOI:** 10.1101/2022.12.22.22283878

**Authors:** Renske Lok, Lara Weed, Joseph Winer, Jamie M. Zeitzer

**Affiliations:** Department of Psychiatry and Behavioral Sciences, Stanford University, Stanford CA 94305; Department of Bioengineering, Stanford University, Stanford CA 94305; Department of Neurology, Stanford University, Stanford CA 94305; Mental Illness Research Education and Clinical Center, VA Palo Alto Health Care System, Palo Alto CA 94304

## Abstract

**Importance:** Human mental and physical health is influenced by both the inclination to sleep at specific times (chronotype) as well as the actual sleep timing (behavior). How the alignment between these impacts mental and physical health has not been well described.

**Objective:** The goal of this study is to examine the impact of chronotype, actual timing of behavior, and the alignment between the two on a variety of mental and physical health outcomes.

**Design:** A cohort analysis of the UK Biobank (2006 to Current).

**Setting:** Outpatient.

**Participants:** Community-dwelling adults (n=73,888).

**Exposure:** *Ad libitum* behavior for one week.

**Main Outcome(s) and Measure(s):** Sleep time preference (chronotype), one week of wrist-worn accelerometry (actigraphy), and demographic variables were collected. Actigraphy was analyzed using non-parametric methods to determine the actual timing of behavior. Prevalence and likelihood (odds-ratios) of developing mental health disorders (mental, behavioral, and neurodevelopmental disorders, generalized anxiety disorder, depression) and physical health disorders (including metabolic disorder, diabetes, obesity, hypertension, circulatory disorder, digestive disorder, respiratory disorder, and all-cause cancer) were calculated and corrected for common demographic variables (sex, age, body mass index, material deprivation, sleep duration).

**Results:** Our final sample was 56% female, 63.5 [56.3 - 68.6] years in age, with a Body Mass Index of 26.0 [23.6 - 29.0], Townsend Deprivation Indices of −2.45 [−3.82 - −0.17], and self-reported sleep duration of 7 [6 - 8] hours. As compared to morning-types with early behavior (aligned), morning-types with late behavior (misaligned) had an increased risk of both mental (OR=1.52±0.06, p<0.001) and physical (OR=1.45±0.03, p<0.001) health disorders. As compared to evening-types with late behavior (aligned), however, evening-types with early behavior (misaligned) had a decreased risk of both mental (OR=0.85±0.06, p=0.002) and physical (OR=0.66±0.03, p<0.001) disorders.

**Conclusion/Relevance:** Despite potential misalignment between sleep and circadian rhythms, going to sleep late is associated with worse mental and physical health in both morning- and evening-types.

## Introduction

The central human circadian clock, located in the hypothalamic suprachiasmatic nucleus (SCN), drives a variety of 24-hour behaviors, predicting changes in the environment and coordinating a myriad of downstream oscillators.^1,2^ One of the most noticeable behaviors under the influence of the SCN is the timing of sleep. While humans are diurnal (day-active), the timing of nocturnal sleep among individuals can vary considerably. Even though the biochemical underpinnings are not well understood, humans often express a preference for going to sleep later (‘owl’) or earlier (‘lark’), a phenomenon known as morningness-eveningness.^3^ This preference has been independently associated with susceptibility to a variety of mental and physical disorders.^4-13^ While humans may prefer a specific time at which to sleep, life may interfere with such plans, causing people to go to sleep later or earlier than they might otherwise prefer. As with morningness-eveningness, going to sleep later and earlier has also been associated with susceptibility to a variety of mental and physical disorders.^14-16^ A smaller number of studies have looked at the synchrony between the preference for sleep timing and the exhibited timing of sleep. When examined, these studies have shown that worse mental and physical health outcomes are associated with such a misalignment.^17-19^ The goal of this study is to examine the impact of morningness-eveningness, actual sleep timing, and the alignment between the two on a variety of health outcomes. To do so, we analyzed data from participants in the UK Biobank, a large, community-dwelling sample of adults.

## Materials and methods

### Ethics

This study was covered by the general ethical approval for UK Biobank studies from the NHS National Research Ethics Service on the 17th of June 2011 (Ref 11/NW/0382). None of the authors had direct contact with the study participants; only deidentified data were used. There were no attempts to identify any of the participants and data were kept secure and only used for the purpose of the approved research, as per the material transfer agreement.^20^

### Accelerometry

Participants used a wrist-worn accelerometer (actigraph: AX3, Axivity, Newcastle upon Tyne, United Kingdom) for one week. Actigraphy files were retrieved from the UK Biobank repository website (5/20/2022). From an initial sample of 103684, 82834 valid accelerometry files were analyzed (Figure 1). Timing of the inactive period was determined by assessing the start time of the five consecutive hours with the lowest level of activity (L5).^21^ Resultant L5 data were categorized in quartiles: Early (Q1), Intermediate (Q2,3), Late (Q4) (Figure 1).

**Figure 1.**
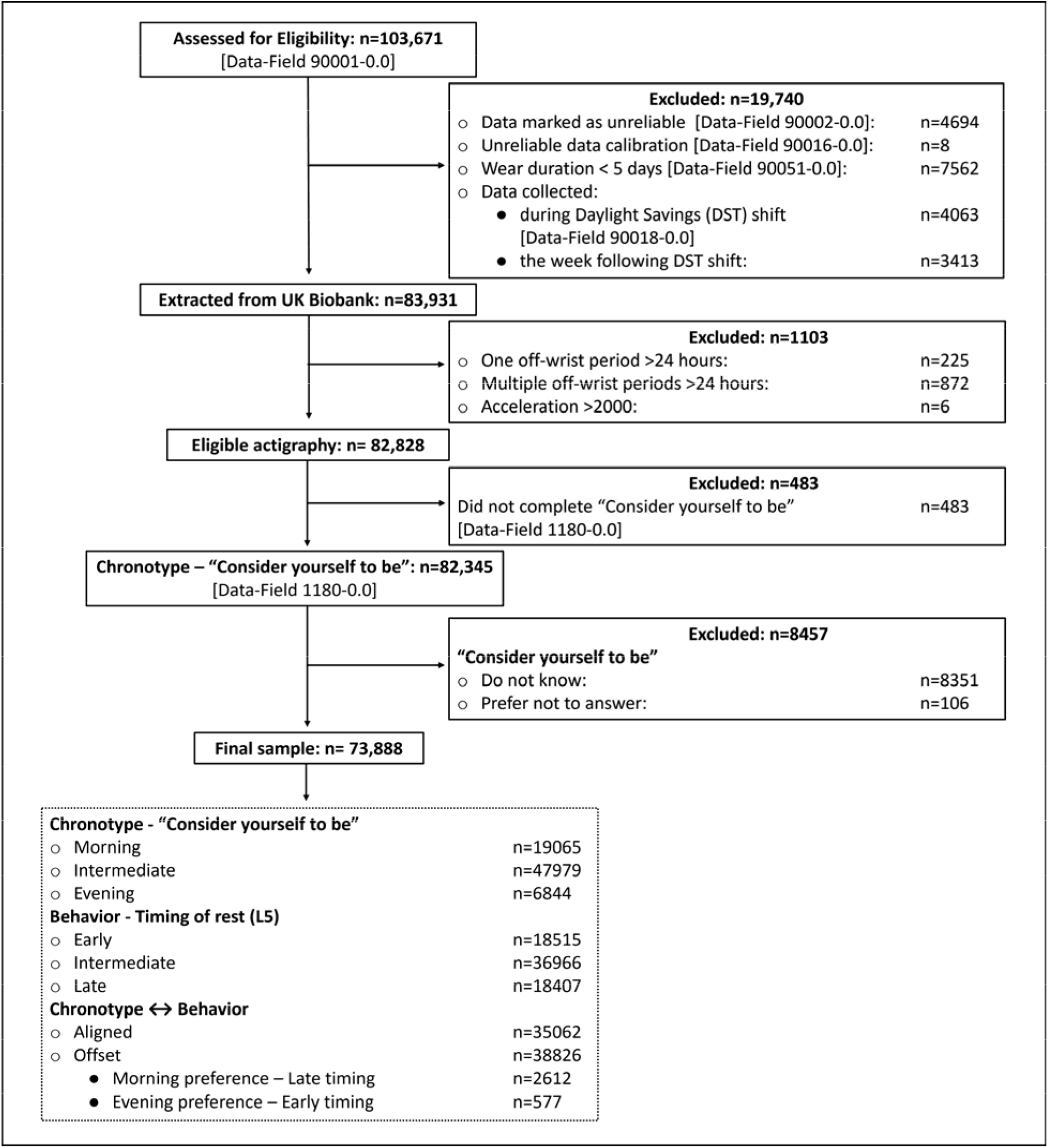
Consort Diagram. In total, 103,671 files were assessed for eligibility and after exclusions, data from 73,888 participants was used. The distribution of both preference for specific times of day (chronotype) and actual timing of behavior (L5) are shown as is the numbers of individuals who are aligned and misaligned between chronotype and L5.

### Chronotype

Morning or evening preference was determined from a single question (n=502413, Data-Field 1180). Individuals responding as “Definitely a ‘morning’ person” were labeled *“Morning*.*”* Individuals responding as “More a ‘morning’ than an ‘evening’ person,” and “More an ‘evening’ than a ‘morning’ person” were categorized as “*Intermediate*.*”* Individuals responding as “Definitely an ‘evening’ person” were labeled as “*Evening”*. Individuals responding as “Do not know” or “Prefer not to answer” were excluded.

### Chronotype↔behavior

To determine the alignment between chronotype and behavior, we compared the quartile overlap in L5 and chronotype. When both L5 and chronotype quartiles were the same, an individual was categorized as “Aligned” (e.g., a morning-type with early behavior). If the L5 and chronotype were misaligned by one step, the individual was categorized as “Offset” (e.g., an evening-type with intermediate behavior). If the misalignment was dichotomous (i.e., chronotype and behavior opposites), an individual was labeled as either “Early Activity ↔ Evening preference” or “Late activity ↔ Morning preference”.

### Mental or physical health disorder diagnosis

To determine the prevalence of mental or physical disorders, the International Classification of Diseases, Tenth Revision, Clinical Modification (ICD10) codes were used (Table 1).

**Table 1.** ICD10 code categorization to determine disorder prevalence.

| Category | Diagnoses | ICD10 codes |
| --- | --- | --- |
| Mental health | Mental, Behavioral and Neurodevelopmental disorders | F01-F99 |
|  | Generalized Anxiety Disorder | F41-1;8;9 <sup>22</sup> |
|  | Depression | F32- A;0;0-5;8;9, F33 <sup>23</sup> |
| Physical health | Metabolic Disorder | E70–E88 |
|  | Diabetes | E10, E11, E13 <sup>24</sup> |
|  | Obesity | E66 <sup>25</sup> |
|  | Hypertension | I10-I15 <sup>26</sup> |
|  | Circulatory Disorder | I00-I99 <sup>27</sup> |
|  | Digestive Disorder | K20-K93 <sup>28</sup> |
|  | Respiratory Disorder | J09-J98, I26-I27 <sup>29</sup> |
|  | All-cause cancer | C0.0-C9.9, D3.7-9, D4.0-8 <sup>29</sup> |

### Statistics

All statistical analyses were conducted in R-studio (version 2021.09.0, running on R version 4.1.2). To test if the observed frequency of disorder diagnosis matched the expected disorder prevalence, or varied due to chronotype, timing of exhibited behavior or the offset between the two, χ tests were used (package stats, version 3.6.2). To determine the likelihood of having a disorder due to chronobiological preference, behavior, or the offset between the two, generalized linear mixed models set for odds ratio (package “oddsratio” and “mfx”) were used. Odds ratios were adjusted for demographic factors, including gender (coded as male/female), age, Body Mass Index (except for the obesity outcome), Townsend Deprivation Index (TDI), and self-reported sleep duration. For all odds ratio tests, the aligned condition functioned as the control (relevel function, package “stats” version 3.6.2). Statistical significance was corrected for multiple testing using a Bonferroni correction;^22^ a p-value <0.0056 was considered significant. Heatmap tables were generated using ggplot (package “ggplot2”). OriginPro 2021b (version: 9.8.5.212, Origin Lab, Northampton MA) was used for all other graphing. Demographic variables are presented as median [interquartile range].

## Results

### General

The UK Biobank includes community-dwelling adults residing in the United Kingdom. Following exclusions for missing data (Figure 1), we had a final sample (n=73888) that was 56% female, 63.5 [56.3 - 68.6] years in age, with a Body Mass Index of 26.0 [23.6 - 29.0] and Townsend Deprivation Indices of −2.45 [−3.82 - −0.17]. Self-reported sleep duration was 7 [6 - 8] hours. Most (97.1%) of the studied population was self-reported to be Caucasian. Given the disproportionate number of Caucasians in the sample, we did not use race as a co-factor in our analyses.

### Chronotype ↔ behavior

Of the final sample, 25.8% were morning-types, 64.9% were intermediate-types, and 9.26% were evening-types (Figure 1). The start of the inactive period (L5) was 00:53 [00:07 - 1:39]. This too was parsed into three categories based on quartiles: early (1^st^ quartile, n=18515, 25.1%, 23:36 [23:13 - 00:12]), intermediate (2^nd^ and 3^rd^ quartiles, n=36966, 50.0%, 00:52 [00:30 - 1:13]) and late (4^th^ quartile, n=18407, 24.9%, 02:08 [1:51 - 2:34]).

Approximately half (47.5%) the sample had alignment between chronotype preference and timing of behavior (Figure 1). Of those with an offset, 3.53% (n=2612) were morning-type with late behavior and 0.78% (n=577) were evening-type with early behavior (Figure 1). Median self-reported sleep duration was 7 hours for all subcategories of chronotype, behavior, and alignment.

### Disorder prevalence – mental health

Mental, behavioral, or neurodevelopmental disorders (MBN, 10.6% of the total included population) were diagnosed in n=7808 participants. Among the MBN were included a diagnosis of Generalized Anxiety Disorder (GAD; n=1921, 2.60%) and depression (n=3248, 4.40%).

Evening chronotype was associated with a significantly elevated frequency of an MBN, including GAD or depression (Figure 2, Table S1). A later timing of behavior was also associated with increased risk for GAD and depression (Figure 2, Table S1). A misalignment between morning chronotype and later timing of behavior was associated with increased MBN prevalence, though these effects were smaller compared to having an evening preference (Figure 2, Table S1). A morning chronotype or exhibiting early to intermediate timing of behavior was somewhat protective against depression diagnosis while exhibiting later timing of behavior was a risk factor (Figure 2, Table S1).

**Figure 2.**
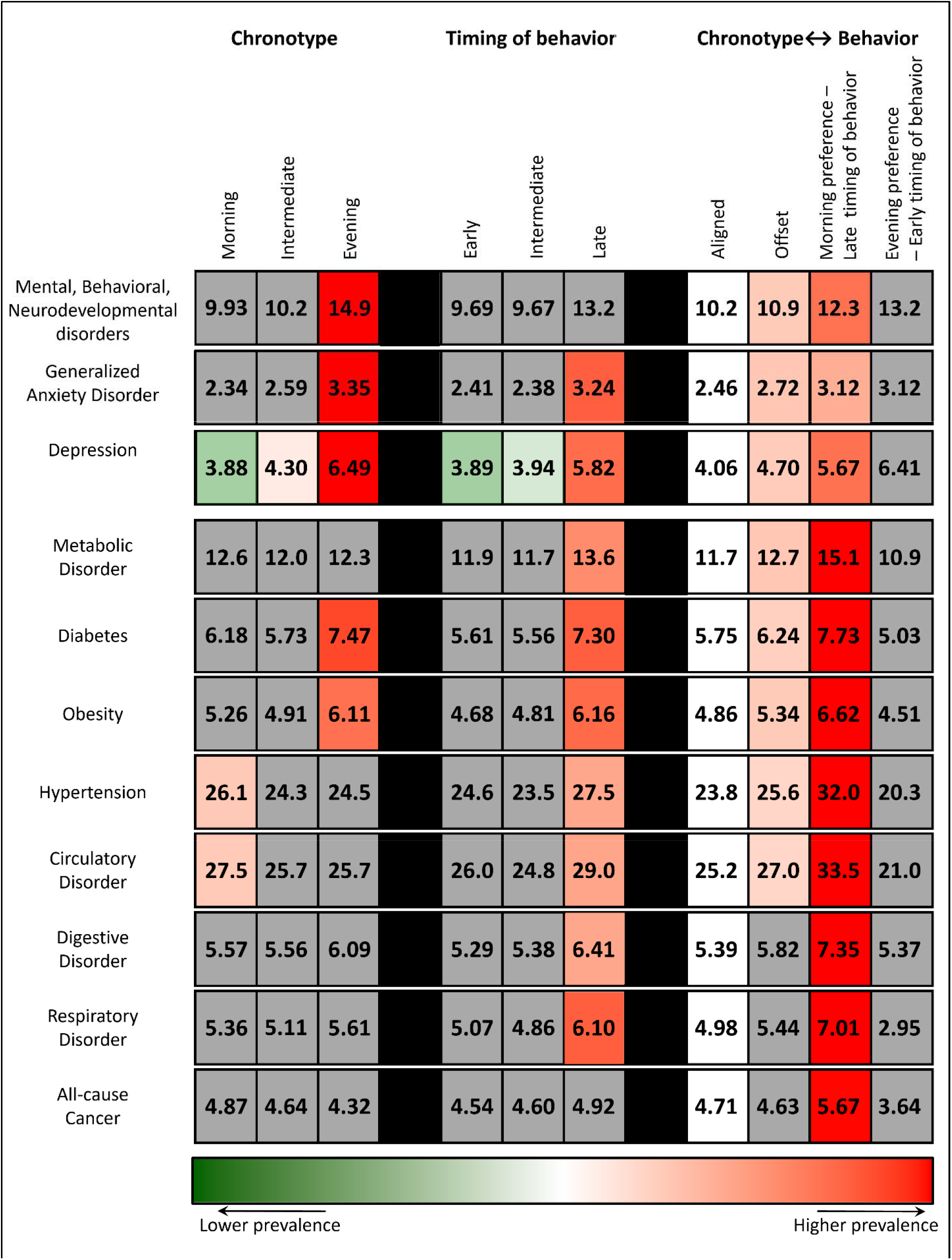
Relative prevalence of selected disorders per chronotype (Morning, Intermediate, Evening), exhibited behavior (timing of behavior categorized as Early, Intermediate, Late) and difference between chronotype and behavior (Aligned, Offset, Late activity with morning preference, or Early activity with evening preference). Individuals having an alignment between chronotype, and behavior functioned as control condition. Non-significant differences between Aligned individuals and other conditions are indicated in grey. Significant increases and decreases in disorder prevalence are presented in red and green, respectively. Color intensity reflects the magnitude of change.

### Disorder prevalence – physical health

Among problems in physical health, there were n=9013 individuals diagnosed with metabolic disorder (12.2%), n=4438 with diabetes (6.01%), n=3777 with obesity (5.11%), n=18290 with hypertension (24.8%), n=19315 with circulatory disorder (26.1%), n=4149 with digestive disorder (5.62%), n=3856 with respiratory disorder (5.22%), and n=3448 with all-cause cancer (4.67%).

More people were diagnosed with diabetes and obesity when having an evening preference. Later timing of behavior was associated with increased diagnoses of metabolic disorder, diabetes, obesity, hypertension, circulatory disease, digestive disorder, and respiratory disorder. In contrast to mental health disorders, physical health disorders were more often diagnosed when there was an offset between chronobiological preference and exhibited timing in rest-activity rhythms (Figure 2, Table S1). The highest prevalence occurred when participants went to sleep late despite having a morning chronotype. For all-cause cancer, this was the only category that had a significantly higher disease prevalence compared to any other category.

### Likelihood of disorder: chronotype

A preference for morning was not associated with changes in the likelihood for diagnosis of MBN, GAD or depression, respiratory disease, diabetes, or all-cause cancer (p’s>0.04), but morning preference was associated with increased likelihood of metabolic disorder, circulatory disorder, or hypertension (p’s<0.001, Figure 3A). Preference for intermediate timing of activity was not associated with a changed likelihood in MBN, GAD or depression (p’s>0.05), nor with diagnosis of metabolic disorder, diabetes, obesity, hypertension, circulatory disorder, digestive disorder, respiratory disorder, and all-cause cancer (p’s>0.05, Figure 3B). However, chronobiological preference for evening was associated with increased likelihood of MBN, GAD and depression, as well as diabetes and obesity (p’s<0.001). Chronobiological preference did not influence likelihood of digestive disorder, respiratory disorder, metabolic disorder, hypertension, circulatory disorder, or all-cause cancer diagnosis (p’s>0.02, Figure 3C).

**Figure 3.**
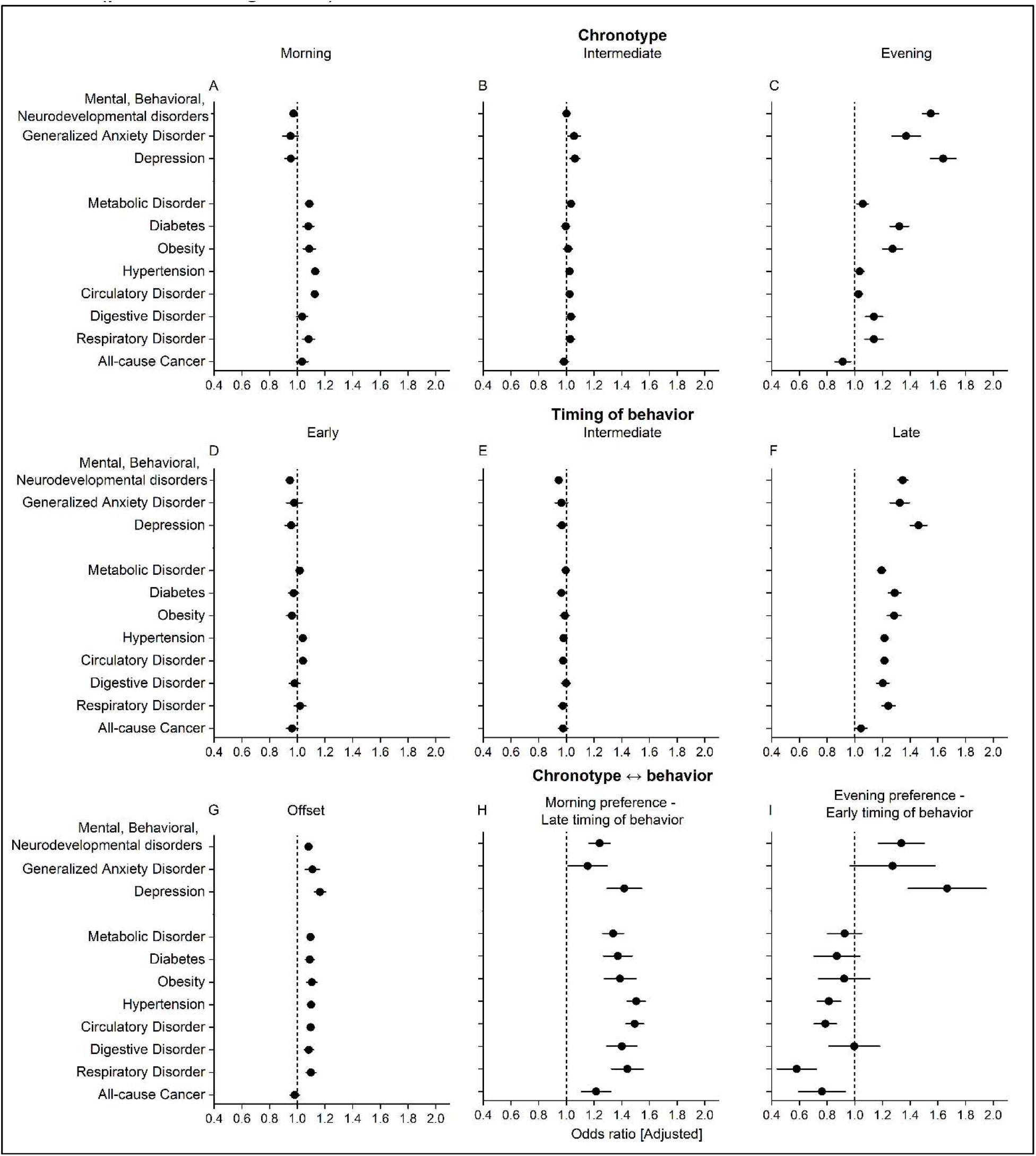
Odds ratio (adjusted for age, gender, body mass index, self-reported sleep duration, and Townsend deprivation index) presented per chronotype (Morning (A), Intermediate (B), Evening (C)), exhibited behavior (timing of behavior categorized as Early (D), Intermediate (E), Late (F)) or the difference between chronotype and behavior (Aligned, Offset (G), Morning preference with late timing of behavior (H), or Evening preference with early timing of behavior (I)). Odds-ratio of 1 (dotted line) indicates no change in likelihood from control condition (match between chronotype and behavior: morning preference with early activity, intermediate preference with intermediate timing of activity, and late preference with late timing of activity). Significant improvements are represented by data to the left of the dotted line, while significant decrements are presented to the right of the dotted line. Odds ratio ± SD are presented.

### Likelihood of disorder: timing of behavior

Displaying an early timing of behavior was not associated with changed likelihood of MBN, GAD, depression, metabolic disorder, circulatory disorder, diabetes, obesity, hypertension, digestive disorder, respiratory disorder, or all-cause cancer (p’s >0.05, Figure 3D). Intermediate timing of rest was also not associated with a significant change in the likelihood of developing any of the included disorders (p’s>0.02, Figure 3E). Late timing of behavior was associated with increased likelihood of MBN, GAD and depression (p’s<0.001), as well as metabolic disorder, diabetes, obesity, hypertension, circulatory disorder, digestive and respiratory disorder (p’s<0.001). It was not associated with a changed likelihood in all-cause cancer diagnoses (p=0.30, Figure 3F).

### Likelihood of disorder: Offset

An offset between preferred and exhibited timing of behavior increased the likelihood of MBN and depression (p’s<0.001), but not GAD (p=0.02). It was also associated with increased likelihood of diagnosis with metabolic disorder (p<0.001), diabetes (p=0.005), obesity (p=0.003), hypertension, circulatory disorder (p’s<0.001), and respiratory disorder (p=0.004). Any offset between preferred and exhibited timing of behavior-activity was not associated with changed likelihood of digestive disorder (p=0.02) or all-cause cancer diagnoses (p=0.59, Figure 3G).

### Likelihood of disorder: Morning preference – late timing of behavior

Preferring morning activity while displaying late timing of behavior increased the likelihood of being diagnosed with MBN and depression (p’s<0.001), but not GAD (p=0.24). Furthermore, it was associated with increased likelihood of being diagnosed with metabolic disorder, diabetes, obesity, respiratory disorder, hypertension, digestive disorder, circulatory disorder (p’s<0.001), but not all-cause cancer (p=0.03, Figure 3H)

### Likelihood of disorder: Evening preference – early timing of behavior

Preferring evening activity while displaying early timing of behavior increased the likelihood of depression (p=0.003), but not MBN or GAD (p’s>0.01). Having this dichotomy was not associated with metabolic disorder, diabetes, obesity, hypertension, digestive disorder, all-cause cancer (p’s>0.05), circulatory disorder, or respiratory disease (p’s=0.02, Figure 3I).

### Likelihood of disorder: Morning preference with early, intermediate, or late timing of behavior

When compared to individuals with a morning preference and early timing of behavior (aligned), individuals with morning preference but intermediate timing of behavior showed no change in the likelihood of developing mental health disorder (p’s>0.02), nor in developing metabolic disorder, diabetes, obesity, digestive disorder, respiratory disorder, or all-cause cancer (p’s>0.04). However, having a morning preference while displaying an intermediate timing of activity significantly decreased the likelihood of developing hypertension or circulatory disorder (p’s<0.001, Figure 4A).

**Figure 4.**
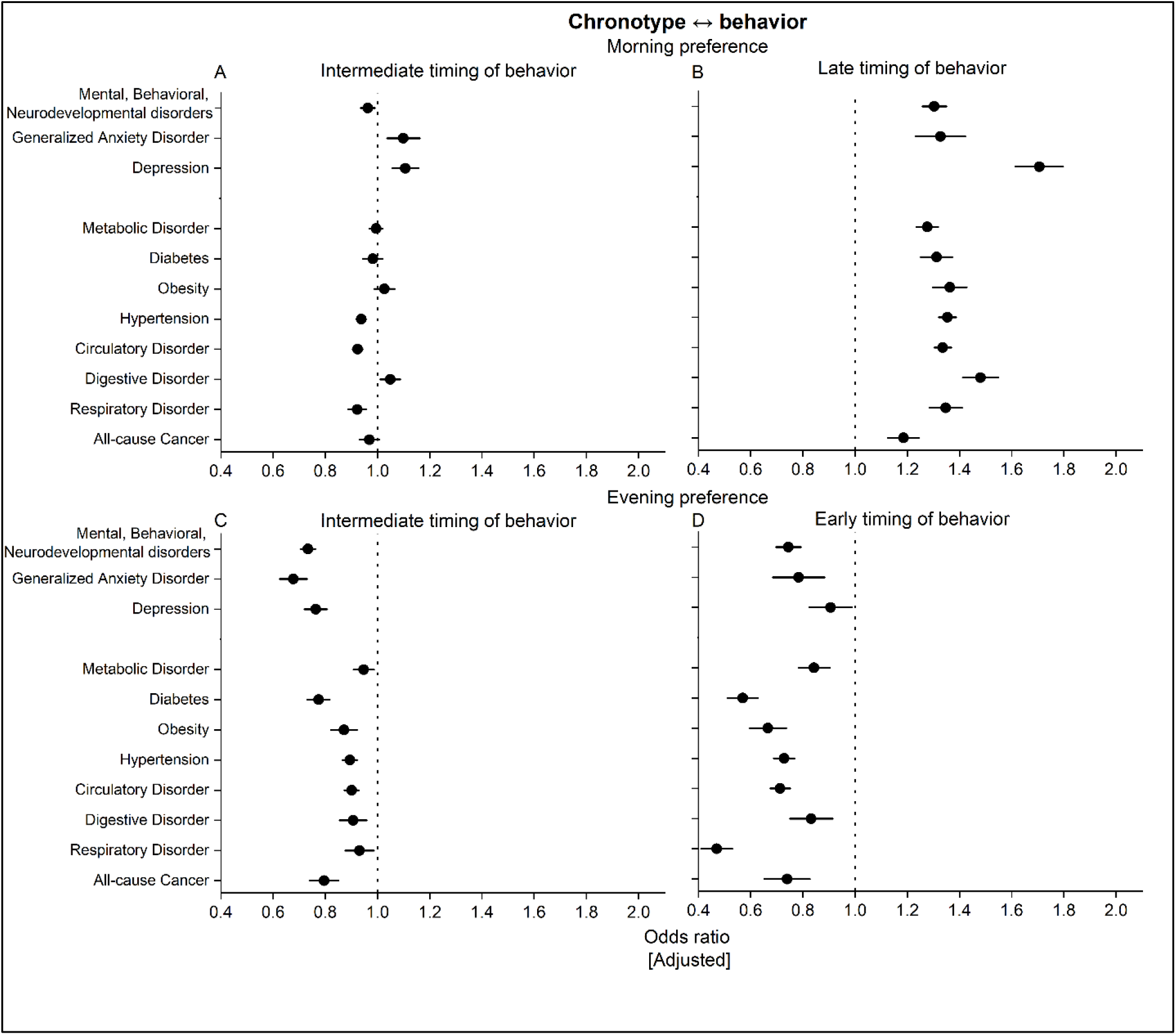
Odds ratio (adjusted for age, gender, body mass index, self-reported sleep duration, and Townsend deprivation index) presented per Morning preference with intermediate (A) or late timing of behavior (B), or Evening preference with intermediate (C) or early timing of behavior (D). Odds-ratio of 1 (dotted line) indicates no change in likelihood from control condition (match between Morning chronotype with Early timing of behavior for the Morning preference group, and Evening preference with late timing of behavior for the Evening preference group). Significant improvements are presented left of the dotted line, while significant decrements are presented to the right of the dotted line. Odds ratio ± SD are presented.

When individuals with a morning preference displayed a late timing of activity, there was a significant increase in the likelihood of having each of the mental health (p’s<0.001) or physical health (p’s<0.001, Figure 4B) disorders that we examined.

### Likelihood of disorder: Evening preference with early, intermediate, or late timing of behavior

When compared to individuals with an evening preference and late timing of behavior (aligned), individuals with evening preference but intermediate timing of behavior showed decreased likelihood of developing all included mental health disorders (p’s <0.001). Being an evening type with intermediate timing of activity also decreased the likelihood of developing diabetes, obesity, hypertension, circulatory disorder, and all-cause cancer (p’s<0.001), while there was no change in the likelihood of developing metabolic disorder, digestive disorder, or respiratory disorder (p’s>0.05, Figure 4C). When individuals with an evening preference displayed an early timing of activity, there was a decreased likelihood of developing MBN (p<0.001), but not GAD (p=0.05) or depression (p>0.05). Furthermore, having an evening preference and displaying an early timing of activity was associated with a decreased likelihood of diabetes, obesity, hypertension, circulatory disorder, respiratory disorder (p’s<0.001), but not metabolic disorder (p=0.01), digestive disorder (p=0.06) or all-cause cancer (p=0.01, Figure 4D).

## Discussion

Having a late schedule, particularly going to sleep after 2AM, is associated with negative mental and physical health, irrespective of chronotype. Alignment of chronotype with the actual timing of activity is beneficial in morning-types as when these individuals are misaligned and keep a late schedule, there is an increased odds ratio for each of the mental and physical health disorders that we examined (Figure 4B). Alignment of chronotype with the actual timing of activity in evening-types, however, is detrimental as when these individuals are misaligned and keep either an intermediate(Figure 4C) or early (Figure 4D) schedule, there is a decreased odds ratio for most of the mental and physical health disorders that we examined.

### Mental health disorders

The association between evening preference and mental health disorders has been previously assessed,^4-13^ and is particularly profound in individuals with depression.^23,24^ Displaying a late timing of activity, predominantly in combination with a morning chronotype, is the biggest risk factor for depression. While underlying mechanisms remain to be elucidated, nocturnal activity is associated with impulsive and maladaptive behavior,^25-27^ and behavioral inhibition declines with prolonged wakefulness.^28^ Functioning of the prefrontal cortex, which is involved in risk assessment, behavioral inhibition, and cognitive control,^29^ is altered during nocturnal wakefulness and could contribute to alterations in mood as well (for a review, see ^26^).

### Physical health disorders

Having a morning preference while exhibiting a late timing of behavior increased the likelihood of having had almost all included physical health disorders. Circadian misalignment, defined as a difference in timing of the endogenous circadian pacemaker and the exhibited sleep-wake pattern, is an important biological component of mood and physiological well-being.^30^ Individuals with a morning preference but late actual timing of activity are typically awake while their circadian clock is signaling for sleep.^31^ This situation is associated with disruptive behaviors,^25-27^ including less nutritious food choices (i.e., skewed toward carbohydrates)^32^ and food intake beyond caloric demands (possibly facilitated by changes in neuroendocrine physiology and hormones),^33^ with the potential for negative health consequences such as obesity, diabetes, impaired glucose tolerance and reduced insulin sensitivity,^34-37^ as well as cardiovascular problems.^34^

Having an evening preference when displaying an early timing of behavior is protective for most physical health disorders. Individuals with this pattern go to sleep during a time at which the circadian alertness signal is elevated, and awaken before their circadian sleep signal has ended.^31^ While awakening at this circadian phase is more difficult, it would provide a more robust light signal to the phase-advance portion of the phase response curve,^38,39^ which may be beneficial for engendering a greater amplitude of the circadian clock.^40^ Enhanced circadian amplitude could contribute to the protective nature of the misalignment of eveningness with an early schedule.

Chronic circadian misalignment, such as occurs in shift workers, increased the risk of breast^41^ and prostate cancer.^42^ In our study, we observe higher prevalence of a history of cancer in morning-type individuals who have a late timing of behavior. While still misaligned, evening-type individuals who have an intermediate or early timing of behavior actually have a lower prevalence of a history of cancer. As with other physical disorders, it may be that the early morning light to which this latter group is exposed may enhance circadian amplitude and be protective against cancer.^40,43-45^ Given the relatively low number of individuals with prostate and breast cancer in this cohort, we were unable to accurately examine whether these specific cancers behaved in a similar or dissimilar fashion to other cancers vis-à-vis synchrony between circadian preference and actual activity timing.

Sleep duration has been much investigated in terms of its relationship to health, with both long (>8 h/d) and short (<7 h/d) self-reported sleep duration being associated with an increased risk of poorer health (for a review, see ^46^). As such, we corrected all odds-ratio models for self-reported sleep duration. Importantly, while our *a priori* assumption was that individuals who slept out of synch with their chronotype would have shorter sleep, we did not observe this. Our data would therefore indicate that the differences in mental and physical health we observe are associated with the offset itself and not with any lack of sleep this offset might cause.

Our study is not without limitations. To correct for difference in sleep duration, we used self-reported sleep duration. While the UK Biobank did not collect electroencephalography,^47^ accelerometry is regularly used to assess sleep and sleep duration. However, accelerometery is known to misestimate sleep durations, predominantly in those who have multiple nocturnal awakenings or take a long time to fall asleep.^48,49^ Various studies have indicated correlations between self-reported sleep duration and long-term health,^46,50-52^ thus our use of this as a correction factor is not wholly unwarranted. To assess disorder prevalence, we relied on ICD-10 codes, which were assessed in different hospitals by different physicians. Though ICD-10 codes are standardized and come with diagnostic guidelines, there may still be inter-physician variation and are unlikely to capture all individuals with a given disorder. Lastly, it must be noted that in this cohort, there were more individuals with a morning than evening preference, while in the general population, eveningness is more prevalent.^53,54^ However, the included population from the UK biobank ranged from 43.5 to 78.9 years, and ageing is associated with earlier chronotypes,^55^ which in part may explain the over-representation of morning chronotypes.

## Conclusion

Humans may prefer to go to sleep at a specific time, but life may interfere. While those who prefer the morning (‘larks’) can go to sleep at early or intermediate times, going to sleep late is associated with worse mental and physical health. Those who prefer the night (‘owls’) have worse mental and physical health when they follow their rhythm predilection and go to sleep late. For these owls, going to sleep at an earlier time is associated with lower rates of mental and physical health disorders. To age healthily, individuals should time their rest before 1 AM, despite their chronobiological preferences.

## Data Availability

All data may be directly accessed through application to the UK Biobank.

## Acknowledgements

This research has been conducted using the UK Biobank Resource under application number 63099. We would like to thank all UK Biobank participants for agreeing to volunteer in this research. Some of the computing for this project was performed on the Sherlock cluster. We would like to thank Stanford University and the Stanford Research Computing Center for providing computational resources and support that contributed to these research results. We would also like to thank Dr. Aly Suh for thought-provoking discussions.

## Author contributions

Dr. R. Lok had full access to all data in the study and takes responsibility for the integrity of the data and the accuracy of the data analysis. Concept and design: Lok, Zeitzer; Acquisition, analysis, or interpretation of data: Lok, Weed, Winer, Zeitzer; Drafting of the manuscript: Lok; Critical revision of the manuscript for important intellectual content: Lok, Winer, Zeitzer; Statistical analysis: Lok; Obtained funding: Zeitzer; Supervision: Zeitzer

## Data Sharing Statement

All data may be directly accessed through application to the UK Biobank.

