## Supplemental Information for "Perils of the nighttime: impact of behavioral timing and preference on mental and physical health in 73,888 community-dwelling adults"

**Affiliations**

**Supplementary materials and methods**

**Description of data.** For a complete description of data collection, see ^21^. Briefly, between February 2013 and December 2015, individuals enrolled in the UK Biobank were invited to wear a wrist-worn triaxial accelerometer (AX3, Axivity, Newcastle upon Tyne, United Kingdom) for seven days. In order of acceptance, participants were sent the accelerometer from June 2013 onwards to January 2016. Participants were informed that the accelerometer should be worn on their dominant wrist continuously, and to start wearing the accelerometer immediately after receiving it. Information regarding demographics, health status, and lifestyle was provided by participants. These measures have been described in more detail elsewhere ^22^.

**Demographic data.** All variables except the Townsend Deprivation Index were collected during the initial Assessment Centre visit. Sex was coded as female/male [Data-Field 31]. Date of birth [Data-Field 33] was used to calculate age at the time of accelerometry collection. Race was collected as part of the touchscreen questionnaire [Data-Field 20001, Data-Coding 1001]. Body mass index was collected in kg/m² [Data-Field 21001]. Average self-reported sleep duration was collected as: “About how many hours sleep do you get in every 24 hours? (please include naps)" [Data-Field 1160, Data-Coding 100291]. If the answer was < 1 or >23 hours, the answer was rejected. If the answer was <3 or >12, the participant was asked to confirm their entry. The Townsend Deprivation Index, a measure of material deprivation in which 0 is the median and positive scores indicate greater deprivation [Data-Field 189], was collected immediately prior to participants joining UK Biobank ^21^.

**Actigraph data preparation.** Data were converted from the original .cwa format to .csv format using the dedicated *biobankAccelerometerAnalysis* package in Python 3 (version 3.1). Data were calibrated to local gravity ^22,23^, in which stationary periods in ten-second windows in which all three axes have a standard deviation of less than 13.0 mg are identified. These stationary periods are then used to optimize the gain and offset for each axis (6 parameters) to fit a unit gravity sphere using ordinary least squares linear regression. Non-wear time was defined as consecutive stationary episodes lasting for at least 60 minutes in which all three axes had a standard deviation of less than 13.0 mg. If present, non-wear segments were automatically imputed using the median of similar time-of-day vector magnitude and intensity distribution data points with one-minute granularity on different days of the measurement ^24^. To optimize data analyses, Sherlock, a high-performance computing cluster provided by Stanford University, was used. Time was coded as 24:00 = 0 and with each hour being a whole number (i.e., an L5 of 02:00 would be coded as 2 and an L5 of 22:00 would be coded as -2).

**Supplementary results.**

Table S1. Results of χ² statistics. Represented are all included disorders (Mental, Behavioral and Neurodevelopmental disorders, Generalized Anxiety Disorder, depression, metabolic disorder, diabetes, obesity, hypertension, circulatory disorder, digestive disorder, respiratory disorder, and all-cause cancer) per category (Chronotype: Morning, Intermediate, Evening; Timing of behavior: Early, Intermediate, Late; the offset between chronotype and timing of behavior; morning chronotype with late timing of activity; and evening chronotype with early timing of activity). Significant values that survived Bonferroni multiple corrections are presented in bold.

| **Disorder** | **Category** | **χ² statistics** | |
| --- | --- | --- | --- |
|  |  | **Degrees of freedom** | **χ², *p*** |
| Mental, Behavioral, and Neurodevelopmental Disorders | Morning chronotype | 1,1894 | 0.650, *p*=0.419 |
|  | Intermediate chronotype | 1,4892 | 0.007, *p*=0.932 |
|  | **Evening chronotype** | **1,1022** | **66.77, *p*<0.001** |
|  | Early timing of behavior | 1,1795 | 2.553, *p*=0.110 |
|  | Intermediate timing of behavior | 1,3575 | 4.213, *p*=0.040 |
|  | **Late timing of behavior** | **1,2438** | **90.21, *p*<0.001** |
|  | **Offset** | **1,4239** | **8.678, *p*=0.003** |
|  | **Morning chronotype, late timing of behavior** | **1,322** | **14.98, *p*<0.001** |
|  | Evening chronotype, early timing of behavior | 1,76 | 4.401, *p*=0.035 |
| Generalized Anxiety Disorder | Morning chronotype | 1,447 | 0.679, *p*=0.410 |
|  | Intermediate chronotype | 1,1245 | 1.388, *p*=0.238 |
|  | **Evening chronotype** | **1,229** | **16.71, *p*<0.001** |
|  | Early timing of behavior | 1,446 | 0.133, *p*=0.714 |
|  | Intermediate timing of behavior | 1,879 | 0.506, *p*=0.476 |
|  | **Late timing of behavior** | **1,596** | **25.91, *p*<0.001** |
|  | Offset | 1,1058 | 4.800, *p*=0.028 |
|  | Morning chronotype, late timing of behavior | 1,74 | 1.313, *p*=0.251 |
|  | Evening chronotype, early timing of behavior | 1,18 | 3.150, *p*=0.075 |
| Depression | **Morning chronotype** | **1,740** | **18.47, *p*<0.001** |
|  | Intermediate chronotype | 1,2064 | 2.678, *p*=0.101 |
|  | **Evening chronotype** | **1,444** | **103.6, *p*<0.001** |
|  | **Early timing of behavior** | **1,720** | **17.76, *p*<0.001** |
|  | **Intermediate timing of behavior** | **1,1456** | **24.17, *p*<0.001** |
|  | **Late timing of behavior** | **1,1072** | **29.66, *p*<0.001** |
|  | **Offset** | **1,1824** | **16.27, *p*<0.001** |
|  | **Morning chronotype, late timing of behavior** | **1,148** | **14.21 *p*<0.001** |
|  | Evening chronotype, early timing of behavior | 1,37 | 7.195, *p*=0.007 |
| Metabolic disorder | Morning chronotype | 1,5775 | 7.310, *p*=0.170 |
|  | **Intermediate chronotype** | **1,148** | **18.82, *p*<0.001** |
|  | **Evening chronotype** | **1,2511** | **33.09, *p*<0.001** |
|  | Early timing of behavior | 1,2197 | 0.302, *p*=0.582 |
|  | Intermediate timing of behavior | 1,4305 | 0.021, *p*=0.884 |
|  | **Late timing of behavior** | **1,2511** | **33.09, *p*<0.001** |
|  | **Offset** | **1,4916** | **12.84, *p*<0.001** |
|  | **Morning chronotype, late timing of behavior** | **1,393** | **20.06, *p*<0.001** |
|  | Evening chronotype, early timing of behavior | 1,63 | 0.257, *p*=0.611 |
| Diabetes | Morning chronotype | 1,1179 | 3.721, *p*=0.053 |
|  | Intermediate chronotype | 1,2748 | 0.016, *p*=0.897 |
|  | **Evening chronotype** | **1,511** | **26.10, *p*<0.001** |
|  | Early timing of behavior | 1,5775 | 0.383, *p*=0.535 |
|  | Intermediate timing of behavior | 1,2055 | 1.095, *p*=0.295 |
|  | **Late timing of behavior** | **1,1344** | **43.31, *p*<0.001** |
|  | **Offset** | **1,2422** | **7.532, *p*=0.004** |
|  | **Morning chronotype, late timing of behavior** | **1,202** | **15.10 *p*<0.001** |
|  | Evening chronotype, early timing of behavior | 1,29 | 0.493, *p*=0.482 |
| Obesity | Morning chronotype | 1,1002 | 3.735, *p*=0.053 |
|  | Intermediate chronotype | 1,2357 | 0.122, *p*=0.727 |
|  | **Evening chronotype** | **1,418** | **16.70, *p*<0.001** |
|  | Early timing of behavior | 1,866 | 0.780, *p*=0.377 |
|  | Intermediate timing of behavior | 1,1778 | 0.079, *p*=0.778 |
|  | **Late timing of behavior** | **1,1133** | **36.28, *p*<0.001** |
|  | **Offset** | **1,2074** | **8.056, *p*=0.004** |
|  | **Morning chronotype, late timing of behavior** | **1,173** | **14.29, *p*<0.001** |
|  | Evening chronotype, early timing of behavior | 1,26 | 0.138, *p*=0.710 |
| Hypertension | **Morning chronotype** | **1,4979** | **21.02, *p*<0.001** |
|  | Intermediate chronotype | 1,11636 | 1.248, *p*=0.264 |
|  | Evening chronotype | 1,1675 | 0.814, *p*=0.366 |
|  | Early timing of behavior | 1,4546 | 2.155, *p*=0.142 |
|  | Intermediate timing of behavior | 1,8674 | 0.792, *p*=0.373 |
|  | **Late timing of behavior** | **1,5070** | **52.71, *p*<0.001** |
|  | **Offset** | **1,9937** | **18.70, *p*<0.001** |
|  | **Morning chronotype, late timing of behavior** | **1,836** | **50.78, *p*<0.001** |
|  | Evening chronotype, early timing of behavior | 1,117 | 2.496, *p*=0.114 |
| Circulatory disorder | **Morning chronotype** | **1,5246** | **20.03, *p*<0.001** |
|  | Intermediate chronotype | 1,12310 | 1.302, *p*=0.253 |
|  | Evening chronotype | 1,1759 | 0.446, *p*=0.504 |
|  | Early timing of behavior | 1,4809 | 2.241, *p*=0.134 |
|  | Intermediate timing of behavior | 1,9160 | 1.037, *p*=0.308 |
|  | **Late timing of behavior** | **1,5346** | **52.52, *p*<0.001** |
|  | **Offset** | **1,10478** | **17.77, *p*<0.001** |
|  | **Morning chronotype, late timing of behavior** | **1,875** | **48.82, *p*<0.001** |
|  | Evening chronotype, early timing of behavior | 1,121 | 3.344, *p*=0.067 |
| Digestive disorder | Morning chronotype | 1,1062 | 0.695, *p*=0.404 |
|  | Intermediate chronotype | 1,2670 | 1.064, *p*=0.302 |
|  | Evening chronotype | 1,417 | 4.843, *p*=0.027 |
|  | Early timing of behavior | 1,980 | 0.203, *p*=0.651 |
|  | Intermediate timing of behavior | 1,1989 | 0.003, *p*=0.195 |
|  | **Late timing of behavior** | **1,1180** | **20.62, *p*<0.001** |
|  | Offset | 1,2259 | 5.687, *p*=0.017 |
|  | **Morning chronotype, late timing of behavior** | **1,192** | **15.76, *p*<0.001** |
|  | Evening chronotype, early timing of behavior | 1,31 | 0.003, *p*=0.985 |
| Respiratory disorder | Morning chronotype | 1,1022 | 3.380, *p*=0.065 |
|  | Intermediate chronotype | 1,2450 | 0.640, *p*=0.423 |
|  | Evening chronotype | 1,384 | 4.292, *p*=0.038 |
|  | Early timing of behavior | 1,939 | 0.206, *p*=0.649 |
|  | Intermediate timing of behavior | 1,1795 | 0.511, *p*=0.474 |
|  | **Late timing of behavior** | **1,1122** | **26.64, *p*<0.001** |
|  | Offset | 1,2111 | 7.107, *p*=0.100 |
|  | **Morning chronotype, late timing of behavior** | **1,183** | **18.30, *p*<0.001** |
|  | Evening chronotype, early timing of behavior | 1,17 | 4.597, *p*=0.032 |
| All-cause Cancer | Morning chronotype | 1,928 | 0.623, *p*=0.429 |
|  | Intermediate chronotype | 1,2422 | 4.533, *p*=0.033 |
|  | Evening chronotype | 1,296 | 1.739, *p*=0.187 |
|  | Early timing of behavior | 1,841 | 0.690, *p*=0.405 |
|  | Intermediate timing of behavior | 1,1702 | 0.4039 *p*=0.525 |
|  | **Late timing of behavior** | **1,148** | **8.436, *p*=0.004** |
|  | Offset | 1,1797 | 0.244, *p*=0.621 |
|  | Morning chronotype, late timing of behavior | 1,148 | 4.418, *p*=0.035 |
|  | Evening chronotype, early timing of behavior | 1,21 | 1.334, *p*=0.247 |
